# Thoughts and perspectives of metagenome sequencing as a diagnostic tool for infectious disease: an interpretive phenomenological study

**DOI:** 10.1101/2024.01.02.24300703

**Authors:** Hannah Trivett, Alistair C. Darby, Oyinlola Oyebode

## Abstract

**Background:** Effective infectious disease diagnostics (IDD) are vital for informing clinical decision-making regarding the treatment and patient management of disease and infections. Conventional clinical methods rely upon culture-dependent techniques, and there has been little shift in the acceptance and integration of culture-independent sequencing methods into routine clinical IDD. This study explored stakeholders’ experiences within IDD, including those working in clinical settings and those conducting research at the forefront of microbial genomics. We aimed to identify factors driving the development and implementation of metagenome sequencing as a routine diagnostic.

**Methods:** Virtual semi-structured interviews were conducted with purposively selected individuals involved in IDD. The interviews explored the experiences of implementing metagenome sequencing as a diagnostic tool and decisions about which diagnostics are used for identifying bacteria-causing infections. Thematic analysis was used to analyse the data, and an Interpretive Phenomenological approach was used throughout.

**Results:** Ten individuals were interviewed between July 2021 and October 2021, including Clinical scientists, consultants, and professors in academia. Their experience ranged from no knowledge of metagenome sequencing to an expert understanding of the phenomenon. Five themes emerged: Diagnostic Choice, Infrastructure, Open Data Sharing, COVID-19, and Communication. Participants recognised the need for new diagnostics to be implemented to overcome the limitations of current diagnostic approaches but highlighted the barriers to integrating new diagnostics into clinical settings, such as the impact on clinical decision-making, accreditation, and cost. However, participants felt that lessons could be learnt from using metagenomics in COVID-19 and how other diagnostic platforms have been integrated into clinical settings over the last 20 years.

**Conclusions:** The study provided clear evidence to address the knowledge gap in current literature and practice for developing and implementing metagenome sequencing as a potential IDD. The knowledge of new and upcoming genomic diagnostic testing is not equally distributed throughout the UK, impacting the understanding and drive to integrate metagenome sequencing into routine clinical diagnostics. Improvements in access to new diagnostics could improve patient treatment and management and positively impact public health.

## Background

The term metagenomics was first published in 1998, referring to directly sequencing a collection of genes from a sample and analysing similarly to a single genome (1). Shotgun metagenomics uses a hypothesis-free, unbiased approach to study the structure and function of the entirety of an environment’s nucleotide sequences, including parasites, fungi, bacteria and viruses. The technological advancements in sequencing have made metagenomic next-generation sequencing (mNGS) an attractive choice for clinical personnel to apply metagenomic methods to infectious disease diagnostics (IDD).

Current clinical diagnostic methods for bacterial pathogens are culture-dependent, requiring the growth and isolation of organisms using methods like serology, Polymerase Chain Reaction (PCR), and Matrix-assisted laser desorption-ionization time of flight mass spectrometry (MALDI-TOF) [2]. However, they are limited by the need for apriori knowledge of the potential pathogens present, and MALDI TOF is limited by the availability of genomes in curated databases to determine the pathogen present within the sample (3). In addition, in some cases, such as those involving slow-growing organisms, it can take as much as two weeks to cultivate a sample, inhibiting the rapid diagnosis and treatment of some infectious diseases.

mNGS can overcome the current limitations of culture-based ‘gold standard’ methods used today in clinical laboratories. Technologies such as Oxford Nanopore offer real-time profiling of bacterial genomes for rapid and accurate pathogen detection. A culture-independent method of surveying the sample site of infection enables clinicians to capture novel pathogens, which may go undetected when using diagnostics such as PCR (4). The cumulation of millions of bacterial sequences provides scopes for mNGS to act as a toolkit identifying pathogens responsible for infections and capturing antimicrobial resistance profiles, functional gene profiling and virulence gene identification simultaneously from a single extracted sample.

Yet, whilst mNGS offers the potential to revolutionise IDD with a suite of clinical and surveillance applications, there has been a shortfall in early adoption to bring this diagnostic into clinical use (5). Why clinicians are not utilising the latest mNGS methods is under-researched, with a lack of qualitative research to understand the current diagnostic landscape and to evaluate why the landscape has remained unchanged for many years. Current literature is saturated with publications on clinical metagenomics, covering topics such as its utility and its limitations when applied to the current diagnostic landscape (6–8); however, there is a gap in the literature which contextualises the recent clinical advancements in IDD and the barriers that hinder the implementation of new diagnostic frameworks. Perspectives and opinions of clinical diagnostic stakeholders influence the uptake of new diagnostic technologies. Therefore, collecting qualitative evidence to understand the utility of mNGS in clinical settings and the barriers to its implementation may allow innovations that will move diagnostics into the next-generation sequencing era.

This study aimed to explore the opinions and views of individuals within IDD on the utility of mNGS to support the future development of genomic diagnostic methods for routine clinical use.

## Methodology

The qualitative enquiry undertook an interpretive phenomenological approach (IPA), documenting the lived experiences of stakeholders participating in the study. IPA is a helpful approach many healthcare researchers adopt to allow others to learn from individuals’ experiences (9,10). The reporting of this study was facilitated by using the consolidated criteria for reporting qualitative research (COREQ) (11).

The study gained ethical approval from the Institute of Life Course and Medical Sciences Research Ethics Committee at the University of Liverpool (Ethical review reference 9855).

### Author reflection

The research team was made up of three individuals, including two women and one man. HT is trained to BSc in Microbiology and is undertaking a PhD in Microbial genomics, supervised by AD and OO, who hold PhDs and are professors in Public Health (OO) and Genomics (AD). HT and OO had training, and OO had previous experience in conducting qualitative data collection. The research team described the reasons for doing the research within participant recruitment emails as being to understand the current landscape of clinical diagnostics for infectious diseases and the added value of clinical metagenomics in clinical laboratories, to support the integration and development of clinical metagenomic pipelines fit for routine clinical use.

### Participant recruitment

Participants were purposively recruited to ensure clinical perspectives that were information-rich with various viewpoints and experiences from the individuals were obtained. Participants had differing levels of epistemology of the phenomenon to ensure a variety of experiences were covered as well as spanning across geographical location, department, length of service and profession (Table 1, Figure 1). The target number of participants for the study was 10. IPA requires a small sample size of around 5-10 participants, allowing for an in-depth analysis of everyone’s experiences (12).

**Figure 1.**
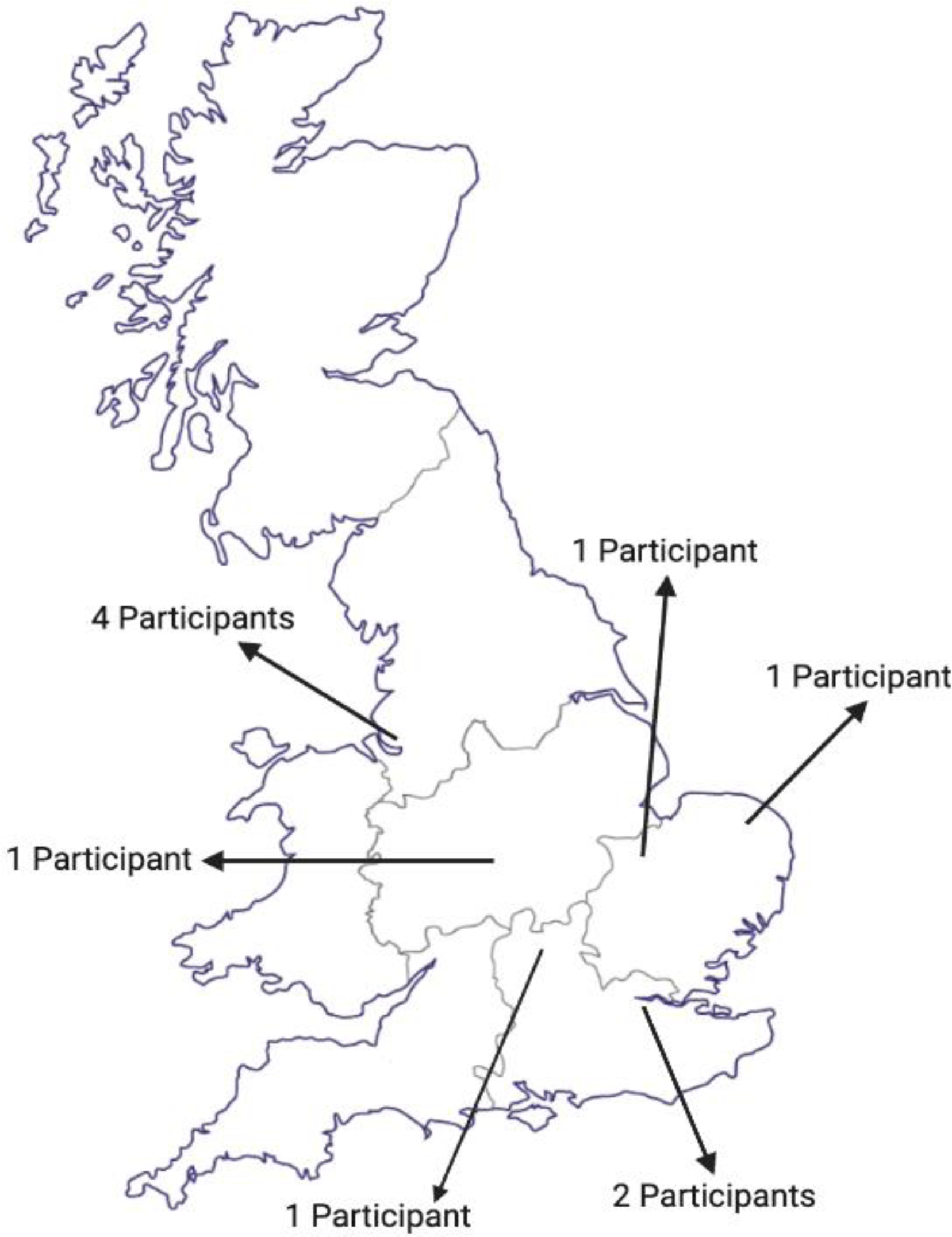
Map of participants’ UK locations. 2 participants are based in London, 1 Participant is based in the Southeast, and 2 participants are in the East of England. One participant was in the West Midlands, and four were in the Northwest.

**Table 1.** Participant demographics.

|  | Gender | Profession | Knowledge of metagenome sequencing applied to Infectious Disease Diagnostics |
| --- | --- | --- | --- |
| **P1** | Female | Consultant clinical scientist and academic researcher | Expert knowledge |
| **P2** | Female | Academic researcher | Expert knowledge |
| **P3** | Male | Infectious disease consultant | Some knowledge |
| **P4** | Male | Consultant and academic researcher | Good Knowledge |
| **P5** | Male | Academic researcher and physician | Expert knowledge |
| **P6** | Male | Consultant clinical scientist | Good Knowledge |
| **P7** | Female | Infectious disease consultant | No Knowledge |
| **P8** | Male | Clinical director | No knowledge |
| **P9** | Female | Academia | Expert knowledge |
| **P10** | Male | Academia | Expert knowledge |

### Data Collection

Semi-structured interviews took place via Microsoft Teams. HT conducted interviews between July 2021 and November 2021. The semi-structured interview guide was created, informed by a literature review, to allow the interviewer to guide the line of questioning—the open-ended questions provided flexibility in exploring topics that may emerge in interviews. The interview guide was checked by two authors (OO and AD) and piloted before beginning data collection with an individual independent of the study. A copy of the guide was sent to participants before their interviews.

### Data analysis

Interviews were video and audio-recorded, and the data were transcribed verbatim in Microsoft Word and analysed using an IPA by HT (2). All data was anonymised, and thematic analysis was conducted. Open coding was performed, identifying experiential statements for each participant, which were then grouped, looking for convergence and divergence between participant experiences; these groups formed the overarching themes of the qualitative enquiry, which were named using words closely linked to the data (13). OO audited the analysis to ensure reliability. A draft of the findings was made available to participants to allow them to check over the study to ensure the information accurately represented their views.

## Results

Ten participants took part in interviews lasting between 30 and 60 minutes. Participant characteristics are shown in Table 1. Five themes were identified from the interviews with all participants, with associated sub-themes (Figure 2).

**Figure 2.**
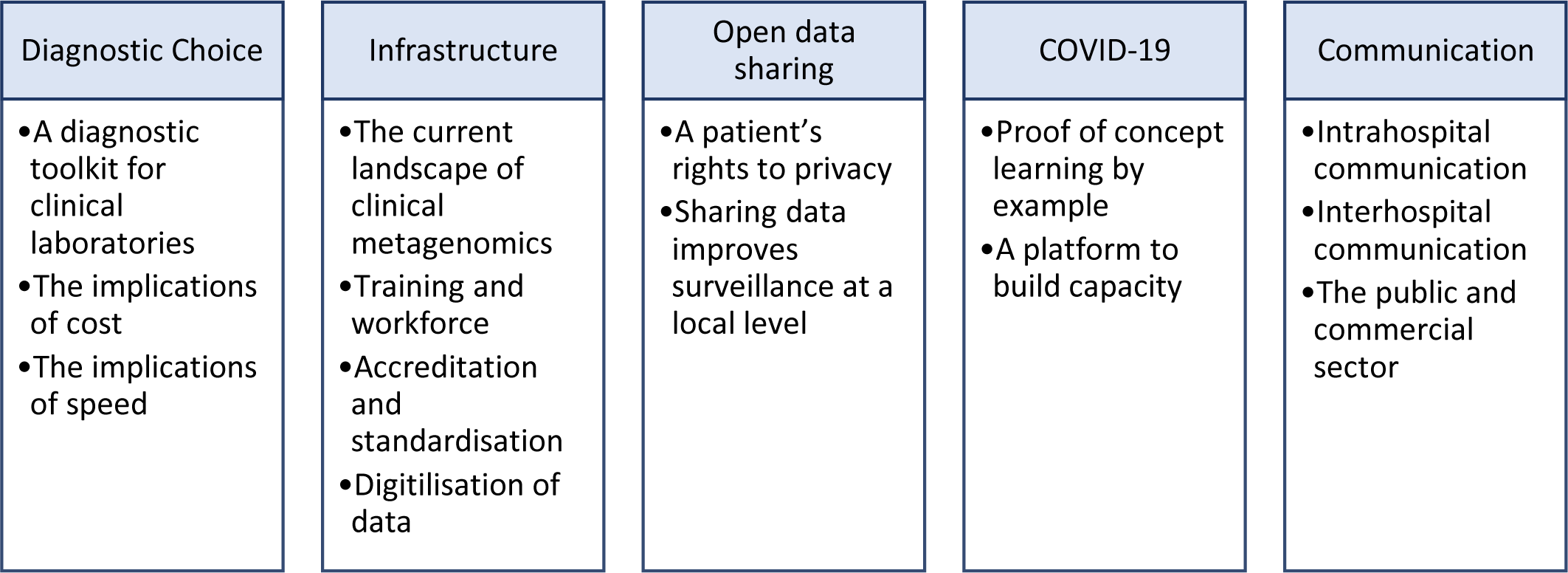
Themes and Subthemes.

### 1. Diagnostic choice

Diagnostic choice was highlighted as an important concept which encompasses the variety of methods to identify pathogens causing infections. Participants offered a range of examples currently available. However, the availability of tools was subjective to each hospital, with some having a whole suite of diagnostics, including genomics, serology, MALDI-TOF and other hospitals limited to culture swabbing to screen for infections and limited PCR.

#### A diagnostic toolkit for clinical laboratories

Throughout the interviews, participants reinforced the benefit of introducing mNGS into clinical laboratories, providing another tool to the armoury and increasing the power of choice to suit clinical needs; however, participants did highlight that mNGS diagnostics would not be a one-size-fits-all approach. Instead, there would be a suitable place and right time to use within the spectrum of tools currently available. Clinical scientists and consultants would have to learn when the appropriate time to use metagenomics to diagnose would be.

*P4 “There are new molecular tests, and actually, there is a steady stream of the introduction of new things to do in response to clinical need. However, there is a set of technologies that are finding it harder to get into the clinical place for infectious diseases, of which genome sequencing is one.”*

*P1 “I don’t think it’s a panacea, I think it’s very much part of our toolkit, and there will be things that are appropriate to use it for, and there will be things that are less appropriate for it to be useful. We have to see it as not one or the other but as a spectrum of clinical choices in terms of diagnostics where, just like everything else, I choose the right test for the right question.”*

#### The implication of cost

There was a broad consensus that cost significantly contributed to the choice of diagnostics on offer, which in some cases impacted the development and integration of metagenomics for routine clinical use. Whilst the affordability of genomic machinery and library preparations has reduced considerably since sequencing first came on the horizon, many commercial providers of diagnostic platforms are still directed towards the research sector, where funding is far more accessible through research grants; thus, distribution is geared towards those who can pay. Several participants stressed that their ability to introduce sequencing into the clinical diagnostic frameworks was due to the procurement of research grants rather than funding available directly from the Trusts or government.

*P2 “All of the sequencing we did was based on a research budget, so we need, we need substantial funding from the NHS in order to roll this out, that’s the first thing.”*

*P1 “If I was working in a normal setting and I wasn’t a researcher, then I would have access to the training, knowledge, understanding and money it takes to bring these things in to get the data that you need to be able to transition it. So, most of our work happens because of the fact that I have research funding.”*

Disparities were seen between smaller district hospitals and those in larger cities with greater access to funding. Participants from smaller hospitals shared their frustrations around the need for more funding to facilitate new diagnostic developments, compared to those from larger city-based hospitals, highlighting the inconsistency in the availability of diagnostics across the UK. Collectively, participants deployed in clinical settings commented on the need for equitable distribution of diagnostics to provide the same level of clinical care for all patients.

*P1 “If you’re going to make a change across the board, we can’t be disadvantaging somebody in South Devon just because of where they live. We need to be doing this in an equitable fashion where those that need it have access.”*

Throughout the interviews, the distribution of money was identified as a critical limiting factor to implementing new diagnostic frameworks, requiring a business case model to reflect the cost-effectiveness and patient benefit of introducing metagenomics. Participants within clinical settings acknowledged the need for better collaboration between those on the laboratory frontline and health economists to bring about a strengthened argument to advocate for the successes of sequencing and the direct impact introducing new diagnostics would have on relieving the patient pathways.

*P8 “It’s been very painful trying to bring those methods on board, and I think it’s been very painful trying to demonstrate the overall cost-effectiveness. All people look at what the acquisition costs are in pathology, and we don’t have enough money, and it’s just taking that whole health economy view, which we are, I think.”*

#### The implication of speed

While cost was a significant barrier to implementing metagenome-based diagnostics, the speed of getting actionable clinical results from metagenomic methods was also a key factor as to why many hospitals do not offer metagenome-based diagnostics in their laboratories. If new technologies can bring about enough change, funders are willing to pay.

*P1 “If it gives you enough of a management change, you can usually find a payoff; what you can’t do is fix time … If I’m not getting it in a clinically actionable time frame, then it is of interest, it is not of use.”*

*P4 “There needs to be a distributed model for the advancement of infectious diseases diagnostic capability, where the testing is done as close to the front line as possible… I think you need to have a more local approach with infectious diseases and testing because things need to be faster, and people need to understand what the clinical implications are.”*

The importance of speed was reiterated when addressing technology’s overall impact on guiding treatment and patient management. Culture-independent methods of diagnosing infectious diseases must reduce the turnaround time between a patient presenting with symptoms and delivering the correct therapy in response to identifying the causative agent. Participants identified that whilst the processing of samples may be relatively short, the time the sample is on the machine may be too long to provide treatment in a clinically actionable time frame, limiting the uptake in implementing metagenomics as a diagnostic.

*P4 “If your result is going to take more than five days to come back, it’s probably irrelevant. I mean, it’s academically interesting, but it’s not going to guide therapy.”*

### 2. Infrastructure

Infrastructure embodies the facilities and systems required to establish metagenomic sequencing in clinical settings. The participant’s perspectives of infrastructure included physical laboratory space, digital space (such as computational setup and storage), and the workforce and training; collectively, all parts of infrastructure must be addressed for the successful integration of metagenomics.

#### Current landscape of clinical metagenomics

Participants reflected on various barriers they experienced, stalling the integration of metagenomic diagnostic frameworks. Although participants could see the potential benefits of introducing point-of-care metagenomics, they commented that they had seen a slow uptake with new technologies, lagging behind use in research-many believed they did not have the infrastructure to facilitate implementation.

*P6 “It’s still so specialised, and should it be? Or should we be able to be offering this as a wider method? And I think, I really think we should be; we just don’t seem to be able to get across that hurdle.”*

Current UK-centralised models of whole genome sequencing and metagenomic analysis delay response time for directed therapies due to the extended turnaround time of shipment, sequencing, and relaying information to the clinical microbiology team. The bottleneck impacts clinicians’ decision-making and could have potential implications of adding to the antimicrobial resistance crisis by administering antibiotics that are not purposefully prescribed for specific bacterial infections. Current responses are primarily to administer empirical treatments before the pathogen is identified. This can prolong the patients’ treatment, widening the gap between admission and treatment with specific narrow-spectrum antibiotics better suited for the infection.

*P7 “I think it’s really important because now we send [a patient sample or isolate] for typing and we get a result, I don’t know, three weeks later, by which time you know you’ve already acted on basis of your clinical suspicion.”*

Decentralising laboratories and providing local infrastructure can overcome many of the obstacles documented by the individuals in the study. One participant described their positive experiences of laboratories in another country where mNGS was well sustained in local hospital laboratories. The infrastructure for mNGS offered solutions to barriers seen in the UK, such as turnaround time when metagenomics and genome sequencing were local to the patients.

*P9 “In Germany, [there are] small regional centres as well as the Robert Koch Institute in Berlin, so they can respond much more quickly in terms of turn-around time because they have all those local labs.”*

#### Training and workforce

Currently, Biomedical and Clinical Scientists have few opportunities to access certified training in bioinformatics, a vital skill for interpreting genomic data to impact clinical decisions for the treatment of patients. Participants indicated that re-training and educating current laboratory personnel is essential for the uptake of new diagnostic frameworks, such as metagenomic sequencing, which is unavailable or has limited availability to clinical staff.

*P9 “I think it would be invaluable for the diagnostics team to have at least some level of [bioinformatic] knowledge because I think that it empowers people also to have the confidence to interpret any data that comes through.”*

Participants based within clinical laboratories expressed that the current array of skills within clinical laboratories lacks the expertise required to establish a full mNGS workflow. Currently, many laboratories rely on local collaborators to interpret bioinformatic data, which has implications for clinicians receiving data within a clinically actionable timeframe. Introducing new team members, such as bioinformaticians, would alleviate the pressures of outsourcing and thus overcome another barrier to implementing metagenomics.

*P3 “One of the things that we are struggling with is that we’re relying on bioinformatic support from elsewhere and not having that on-site.”*

However, several individuals acknowledged that workforce staffing levels could be better and that introducing new metagenomic diagnostics frameworks may be beyond the current scope of these staff numbers.

*P6 “If we are introducing an entirely new workflow, it has a staffing implication and generally, labs are poorly staffed at the moment.”*

#### Accreditation and standardisation

Participants working within microbial laboratories shared the constraints they have experienced when bringing new diagnostic techniques to test, validate and roll out. Many reagents are labelled as ‘research use only’, which restricts clinical usage as the reagents do not qualify for medical application, limiting the availability of reagents that are permitted for medical purposes.

*P1 “Accreditation is a nightmare, so all of our tests have to be accredited, so they have to be ISO accredited by a group called UKAS who come in and assess us. All of the reagents that are used for whole genome sequencing and metagenomics say they’re for research use only, and therefore, to get accreditation with a whole bunch of research reagents is incredibly difficult.”*

In addition, providing evidence for the standardisation of diagnostics processes could be improved. One individual highlighted that many clinical samples are small in quantity, meaning processes cannot be standardised with the same sample; data would be varied with no standard control to validate against. The metagenomic pipeline for diagnostics is far-reaching, applying to various sample types and microorganisms; however, participants identified this as challenging. The variety of outcomes from analyses could complicate the standardisation processes as each sample would, in principle, need its validation.

*P10 “I think you could validate metagenomics diagnostic service, but the outcome would probably not be a diagnosis. The outcome would probably be a data set, you know, because it would be hard to put in a random sample, and you come out with an answer because we just don’t have enough.”*

#### Digitalisation of data

During the interview process, there were frequent discussions of inadequate internal laboratory systems used for data storage for storing large data sets produced by genomic machinery. Many organisations have begun to digitalise their data storage and management system infrastructure to access patient data, streamlining data accessibility. With extensive data footprints in genomics, IDD departments should engage with other departments to learn from others’ experiences in digitalisation to help integrate and develop genomics in healthcare.

*P8 “This sort of data won’t fit into most laboratory systems, I would suggest. So, there’s a lot of work ongoing at the moment about digitising cellular pathology, and you might want to piggyback onto that to find ways of getting this data stored.”*

The infrastructure required for the technology spans the whole pipeline from physical laboratory space to data storage to technology for clinical interpretation. Participants questioned their ability to understand and clinically interpret the data. Participants flagged the need for infrastructure that is easy for individuals to get to grips with and that can provide the correct information that is clinically usable, not just laboratory infrastructure and data storage.

*P7 “I think all technology has to be right, not just the assay, but the reporting side of things. But we need kind of more insight as to how to report them in a clinically useful way.”*

### 3. Open data sharing

Genomic data is often open access and widely shared within the research community. Having data openly available and easy to access accelerates research to find new ways of testing new hypotheses and developing new analysis methods. Participants recognised that challenges are met by the ethical considerations of a patient’s right to privacy versus sharing healthcare data to learn and develop genomic expertise for wider public health benefit.

#### A patients’ rights to privacy

Open accessibility and the ability to share data are core principles for developing and understanding infectious diseases. However, patient information from clinical records is confidential, and restraints are in place to maintain patient privacy of personal information. Here, we see a conflict between the core principle of healthcare systems respecting a patient’s right to privacy and the ability to expand our public data repositories to keep up with evolving infectious diseases and knowledge of diseases.

*P10 “Genomics comes from an open data kind of culture, but clinical diagnostics most certainly doesn’t for obvious reasons, and so does public health epidemiology. So, there’s a culture clash there between whether genomes are public goods, which should be deposited in the public database as soon as you get them, versus people’s rights to privacy for their diagnosis, and this is the general culture of public health which is more closed which may need changing a little bit.”*

#### Sharing data improves surveillance at a local level

A comprehensive application of data sharing impacts individual patients for diagnostic purposes and disseminates knowledge and understanding of the disease and epidemiology, guiding therapeutic development. Participants highlighted the positive impact of a local data generation fed into a centralised data repository. This data can guide disease surveillance and be used to understand better infectious diseases and their evolution, similar to what was seen from COG-UK and the developments in the knowledge of COVID-19.

*P2 “Data generated locally can be collected for national surveillance, and that’s an amazing opportunity…. If you had a centralised database, you could get some amazing insights into what’s happening across the country.”*

### 4. COVID-19

COVID-19 was one of the largest modern-day international outbreaks documented, and the use of genomics to identify and survey the virus is a prime example of the utility of metagenomics within public health. Collectively, participants commended the use of metagenomics in this outbreak, offering several benefits they saw from its implementation worldwide.

#### Proof of concept, learning by example

Participants offered an insight into their experience and opinions of the capabilities of utilising genomics. They believed the COVID-19 model provided proof of concept for further integration as a diagnostic in a clinical setting. COVID-19 provided evidence for rapid delivery of sequencing and its positive impact on public health.

*P2 “SARS-CoV-2 is an exemplary example of where sequencing can bring impact to public health and individuals, so it’s gone beyond proof. It is being used every day and based on that, and I don’t think there’s any going back to thinking that sequencing is some sort of luxury, and so now we need to work out where it’s going to be used when it’s going to be used.”*

Participants highlighted that in recent years, there had been an uptake in routine whole genome sequencing for several pathogens within the centralised model of the genome sequencing service provision provided in the UK, including Tuberculosis and Salmonella. Most recently, metagenomics was used to identify COVID-19, providing an excellent example of the positive impact metagenome sequencing can have on public health.

*P10 “The COVID experience, I think, pushed the argument forward for routine use of sequencing, as nothing else has done. There are routine uses of genomics now for TB, Gram-negative foodborne pathogens like salmonella, HIV, hepatitis maybe, but those are the exceptions rather than the rule of thumb.”*

#### A platform to build capacity

The influx in demand for sequencing technology and reagents from COVID-19 sequencing meant that an increase in manufacturing capacity was established. Increased production of reagents and sequencing platforms has improved the accessibility of consumables to medical laboratories, increasing the likelihood of integrating new diagnostic frameworks into IDD departments.

*P1“The big advantage of all the stuff that’s going on for me in the whole genome sequencing world is that it is driving manufacturers to make stuff that is useful for me. So even though I’m not getting the data, the benefit will be that I now have like the next evolution of Nanopore which means that I can run one or two samples instead.”*

Participants expressed positivity towards what COVID-19 sequencing platforms had done for the visibility of genomic sequencing, pushing forward the case for implementing metagenome sequencing as a routine clinical diagnostic.

*P5 “The pandemic and the experiences with sequencing SARS-CoV-2 bought it into very sharp focus, and that it has motivated not only governments and health agencies but indeed has started to resonate with the public.”*

### 5. Communication

Participants collectively acknowledged communication was a driver for implementing mNGS diagnostic frameworks in clinical laboratories, identifying three key relationships that facilitate this: communication channels within hospitals, between hospitals, and between hospitals and other organisations, such as academia and commercial businesses.

#### Intrahospital communication

Intrahospital communication is defined as the sharing of information within one institution. UK hospital organisation sees departments running independently of one another, with pots of money funding departments in silos and not equal between different disciplines. With departments working autonomously, there needs to be more communication between groups, which can make healthcare disjointed.

*P9 “Hospitals are really bad at speaking between disciplines in different clinical care.”*

One participant compared their experiences of working in UK and German hospitals. Communication channels within hospital departments were more established, with crosstalk between departments, utilising genomic machinery that other departments already have in play. Collaborating within organisations provided efficiency in integrating new technologies, rapidly adopting new diagnostic workflows, and positively impacting patient management and treatment with the latest technology.

*P9 “I walked around the big paediatric hospital here in [German city], so this was with the infectious disease paediatrician, he took me to the guys that all do the rare diseases, and he goes, ‘we just use all of their Novaseq to do our sequencing. Why would we buy it when they’ve got it down the corridor?’ and I’m like, 100%, but I was like a bit like, you know, mind blown because I was like, this doesn’t necessarily happen in other situations.”*

#### Interhospital communication

Interhospital communication is the sharing of information between multiple sites of clinical organisations. Participants highlighted that early adoption of metagenome sequencing will likely be in larger hospitals. The knowledge and experience of setting up and establishing frameworks should be disseminated to other sites to streamline the integration of metagenomics for other organisations.

*P4 “Other places, each one will have their journey, and so it just needs to be focused on. We need to learn from a few sites that have started to do it and listen to them like the conversation we’re having now and share experiences.”*

However, other participants believed not all hospitals easily communicate their experiences to other microbiology diagnostic teams to help others transition to new mNGS clinical diagnostics.

*P1 “I just get really concerned that all of these things happen in pockets and by people do stuff in pockets without a strategic view, without them feeding back to the right places. Then actually, it’s just all of us repeating work.”*

A few participants focused on their positive collaboration experience during COVID-19, where effective communication between local hospitals and COVID-19 sequencing hubs improved collaborative efforts. This streamlines responses, the use of genome sequencing, and learning from the experiences of others to enhance the integration of these technologies.

*P6 “I think it’s been really helpful, particularly in our network, there was the weekly meetings have been very useful for disseminating information about what we need because there was so much change happening; being able to deal with that as a group is better than everyone just doing their own thing and essentially duplicating work to deal with problems.”*

#### The public and commercial sector

Involving all participants in implementing new diagnostics for clinical use is vital for innovation. Participants collectively commented on better communication between all parties engaged in clinical diagnostics.

*P9 “Definitely a disconnect between the scientists that are maybe trying to drive this forward versus the people that are actually in the clinic. I think improving that communication between them is really important when we do this.”*

Participants within clinical settings highlighted that metagenomics is not the first technology system integrated into clinical settings, calling on their experience of MALDI-TOF integration into clinical laboratories. Integration and the development of new technologies require clear communication channels to produce clinically relevant platforms and solve current problems microbial laboratories see today in diagnostics. Improving communication between all parties, academia, industry, and healthcare, would increase productivity in developing clinically relevant products that are fit for purpose.

*P8 “Working with manufacturers is important, isn’t it. because increasingly, we’re seeing the big manufacturers trying to set up whole systems for laboratories that integrate. So, there’s a blood culture machine, there’s a MALDI machine, and there’s automatic sensitivity testing, there’s maybe some PCR machines. So, it’s getting those manufacturers to develop the platforms to integrate metagenomics into the whole laboratory and that that would help move things forward as well.”*

*P4 “You need to align the funding bodies. We need to bring in all the stakeholders as equal partners…I think it needs an insightful, joined up, sort of mutually supporting and engaging partnership framework so that everyone can benefit from it.”*

## Discussion

The study aimed to assess the opinions and perspectives of stakeholders of IDD, using their experiences to understand the current diagnostic landscape of infectious disease and the implications affecting the integration of metagenomics for routine clinical use. To the best of our knowledge, the findings outlined in this paper contribute to the first interpretative phenomenological account of experiences surrounding this topic.

From the analysis, it was clear that the reflections from participants presented a complexity of interrelated components that fell into categories of barriers and/or facilitators that impact the implementation of metagenomics as a routine clinical diagnostic (Figure 3). Almost all participants shared similar perspectives on what they identified as barriers to implementing metagenomics in clinical settings. However, the extent of the impact of these barriers was subjective to the location of participants. These barriers directly correlate with the availability of funds, infrastructure, and relations with local research groups, universities, and commercial partners.

**Figure 3.**
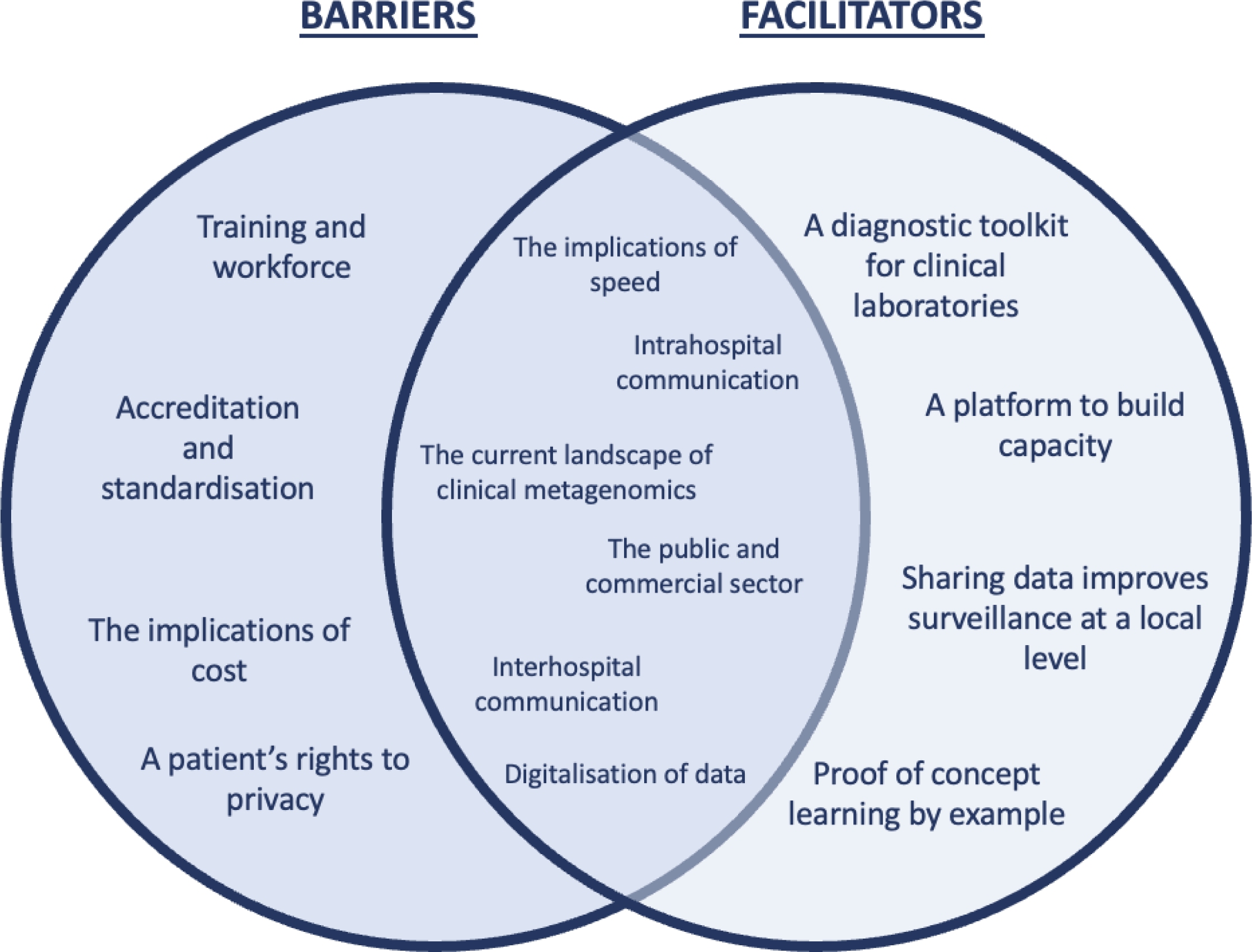
Schematic partitioning of subthemes as barriers and/or facilitators to implementing metagenomics as a clinical diagnostic in infectious disease laboratories.

The qualitative data was supported by many of the current findings of literature reviews, which discuss similar barriers to metagenomics in clinical laboratories, including lack of training and expertise in the workforce, validation of workflows and lack of physical and digital space (4,14). There is a lack of a systematic approach to help level up IDD suites in clinical settings, which has impacted the uptake of routine mNGS at the local level. Many hospitals participating in pathogen sequencing rely heavily on external laboratories to carry out the workflow. With speed identified as a key barrier for implementing new diagnostics, transitioning away from centralised laboratories and moving sequencing to the source hospital could provide an opportunity to improve the speed of the pathogen identification (15). For cases where mNGS may not be local to the hospital, the practicalities of sending samples away for sequencing and waiting for results to be analysed outside of local hospital infrastructure can make mNGS inefficient in its current form as an external diagnostic. Removing time-consuming processes like preparing samples to be sent to external laboratories, mNGS could be faster than conventional culture-dependent bacterial identification. Offering a rapid turnaround from sample collection at the patient bedside to pathogen identification in less than 24 hours would benefit public health through rapid diagnosis to inform patient treatment and public health management (16–18). When developing new diagnostic methods, the whole workflow must be considered to ensure that methods are fit for purpose. Whilst a diagnostic workflow in research may be faster than current clinical methods available, the difference in infrastructure between research and clinical settings may prove the technology to be inefficient in practice, which could hinder the transition of diagnostic methods into clinical laboratories. Therefore, it is necessary to consider what current infrastructure is available on site and what may be logical and practical investments to facilitate the implementation of new diagnostics.

Despite the perceived barriers faced in hospitals delaying metagenomic implementation, it is worth noting that participants were enthusiastic about the potential of new diagnostics in laboratories to be integrated into the suite of tools already offered for infectious diseases. While COVID-19 was challenging for researchers and clinicians, the pandemic positively reinforced the benefits of utilising metagenomics to identify and monitor infectious diseases. High-profile outbreaks and infectious disease events result in an influx of funding and expedite the turnaround time for receiving accreditation, which allows for fast implementation of new technology in clinical settings. Applying metagenomics to clinical samples enables the identification of rare or unknown agents of infectious disease (19,20). In 2022, the UK government implemented a plan for integrating genomics into mainstream healthcare. It followed the 2020 Genome UK: The Future of Healthcare, which outlined an approach to becoming a world leader in the genomic healthcare (21). By bringing genomics to the forefront of diagnostics in healthcare, there is the opportunity to build capacity for the routine use of metagenomics in infectious disease, which will improve guidance for patient treatment and facilitate disease surveillance that is not already part of standard surveillance programmes (22).

mNGS has shown great potential for the detection of pathogens of infectious diseases, providing a sensitive method that can profile taxonomy, antimicrobial resistance and virulence through several clinical pilot studies and case reports (23–25). However, evidence of the cost-effectiveness is also required to provide an argument for its added value to clinical laboratories. The operational value must be presented to inform clinical management of the benefit of technologies such as mNGS (6). Evidence must be provided to highlight the method’s performance compared to currently available diagnostics and its impact on clinical decision-making. The cost-effectiveness of mNGS could be argued through a cost-effective analysis, which would consider not only the economic benefit of mNGS but its public health impact through Quality Adjusted Life Years and an overall number of infections, similar to that published by Elliot et al. (26).

### Strength and limitations

This study is the first of its kind to explore the perspectives of individuals involved with clinical metagenomics as an infectious disease diagnostic. Recruitment took place in various locations across England, covering six regions, and included several stakeholders, allowing us to explore the varying perspectives across diverse professions associated with IDD. However, the Southwest and Northeast of England did not have any representation among the participants. Whilst several drivers are key defining features found across all interviews, without representation, it is unknown if the findings apply to England’s two regions without representation. One of the key strengths was the diversity in expertise across the researchers, spanning public health policy and genomics in both research and clinical settings, which ensured the question guide was reviewed from various perspectives.

## Conclusions

The study filled a gap in the literature around what is known about the development and implementation of metagenome sequencing for routine clinical use. It provided clear guidance for user requirements from new diagnostic technologies to incorporate into the current diagnostic landscape. Participants’ experiences of existing diagnostic workflows and the bottlenecks within them highlighted key drivers that must be considered when developing new diagnostics fit for purpose, such as cost, speed, and infrastructure. High-profile outbreaks such as COVID-19 showcase the utility of metagenomics as a diagnostic tool to benefit public health, which should be used as a learning resource to help innovate IDD with mNGS to bring clinical microbiology into the next-generation sequencing era.

## Supporting information

supplementary COREQ checklist for reporting

## Data Availability

The dataset (which includes individual transcripts) is not publicly available due to confidentiality.

## List of abbreviations

IDD: Infectious disease diagnostics
mNGS: Metagenomic next-generation sequencing
PCR: Polymerase Chain Reaction
MALDI-TOF: Matrix-assisted laser desorption-ionization time of flight mass spectrometry
IPA: Interpretive phenomenological approach
COREQ: Consolidated criteria for reporting qualitative research

## Author declarations

### • Ethics approval and consent to participate

The study gained ethical approval from the Institute of Life Course and Medical Sciences Research Ethics Committee at the University of Liverpool (Ethical review reference 9855). All methods were carried out following relevant guidelines. All participants were provided with an information sheet and provided written consent before participating in this study.

### • Consent for publication

Written consent at the beginning of the project included taking part in the study and consent to anonymised quotes to be used in publications.

### • Authors’ contributions

HT, OO and AD designed the study. HT collected, and HT and OO analysed the data. HT drafted the manuscript and final version, OO and AD contributed to the writing. All authors approved of the final manuscript. Funding was secured by AD from NIHR.

## • Acknowledgements

With thanks to Steve Paterson for help trialling the study guide, along with thanks to Sophie Staniszewska for support during the initial development of this project.

