## supplementary COREQ checklist for reporting for "Thoughts and perspectives of metagenome sequencing as a diagnostic tool for infectious disease: an interpretive phenomenological study"

### Consolidated criteria for reporting qualitative studies (COREQ): 32-item checklist

| No. Item | Guide questions/description | Reported on Page # |
|---|---|---|
| **Domain 1: Research team and reflexivity** | | |
| *Personal Characteristics* | | |
| 1. Inter viewer/facilitator | Which author/s conducted the interview or focus group? | 8 |
| 2. Credentials | What were the researcher's credentials? E.g. PhD, MD | 5 |
| 3. Occupation | What was their occupation at the time of the study? | 5 |
| 4. Gender | Was the researcher male or female? | 5 |
| 5. Experience and training | What experience or training did the researcher have? | 5 |
| *Relationship with participants* | | |
| 6. Relationship established | Was a relationship established prior to study commencement? | n/a |
| 7. Participant knowledge of the interviewer | What did the participants know about the researcher? e.g. personal goals, reasons for doing the research | 5 |
| 8. Interviewer characteristics | What characteristics were reported about the inter viewer/facilitator? e.g. Bias, assumptions, reasons and interests in the research topic | 5 |

| | | |
|---|---|---|
| **Domain 2: study design** | | |
| *Theoretical framework* | | |
| 9. Methodological orientation and Theory | What methodological orientation was stated to underpin the study? e.g. grounded theory, discourse analysis, ethnography, phenomenology, content analysis | 5 |
| *Participant selection* | | |
| 10. Sampling | How were participants selected? e.g. purposive, convenience, consecutive, snowball | 5 |
| 11. Method of approach | How were participants approached? e.g. face-to-face, telephone, mail, email | 8 |
| 12. Sample size | How many participants were in the study? | 6 |
| 13. Non-participation | How many people refused to participate or dropped out? Reasons? | n/a |
| *Setting* | | |
| 14. Setting of data collection | Where was the data collected? e.g. home, clinic, workplace | 8 |
| 15. Presence of non-participants | Was anyone else present besides the participants and researchers? | n/a |
| 16. Description of sample | What are the important characteristics of the sample? e.g. demographic data, date | 5-7 |
| *Data collection* | | |
| 17. Interview guide | Were questions, prompts, guides provided by the authors? Was it pilot tested? | 8 |
| 18. Repeat interviews | Were repeat inter views carried out? If yes, how many? | n/a |
| 19. Audio/visual recording | Did the research use audio or visual recording to collect the data? | 8 |
| 20. Field notes | Were field notes made during and/or after the interview or focus group? | n/a |
| 21. Duration | What was the duration of the inter views or focus group? | 9 |
| 22. Data saturation | Was data saturation discussed? | n/a |
| 23. Transcripts returned | Were transcripts returned to participants | 8 |

| | | |
|---|---|---|
| | for comment and/or correction? | |
| **Domain 3: analysis and findings** | | |
| *Data analysis* | | |
| 24. Number of data coders | How many data coders coded the data? | 8 |
| 25. Description of the coding tree | Did authors provide a description of the coding tree? | n/a |
| 26. Derivation of themes | Were themes identified in advance or derived from the data? | 9 |
| 27. Software | What software, if applicable, was used to manage the data? | n/a |
| 28. Participant checking | Did participants provide feedback on the findings? | 8 |
| *Reporting* | | |
| 29. Quotations presented | Were participant quotations presented to illustrate the themes/findings? Was each quotation identified? e.g. participant number | 5-24 methods and results |
| 30. Data and findings consistent | Was there consistency between the data presented and the findings? | 25-29 discussion |
| 31. Clarity of major themes | Were major themes clearly presented in the findings? | 9 |
| 32. Clarity of minor themes | Is there a description of diverse cases or discussion of minor themes? | 19-29 results and discussion |
